# Asthma in COVID-19: An extra chain fitting around the neck?

**DOI:** 10.1101/2020.07.13.20153130

**Authors:** Mohammad Hosny Hussein, Eman Ali Toraih, Abdallah S Attia, Mohanad Youssef, Mahmoud Omar, Nicholas Burley, Allen D. Zhang, Jackson Roos, August Houghton, Nedum Aniemeka, Mohamed Ahmed Shama, Juan Duchesne, Emad Kandil

## Abstract

**Introduction:** The novel coronavirus disease 2019 (COVID-19) has rapidly spread across the globe, overwhelming healthcare systems and depleting resources. The infection has a wide spectrum of presentations, and pre-existing comorbidities have been found to have a dramatic effect on the disease course and prognosis. We sought to analyze the effect of asthma on the disease progression and outcomes of COVID-19 patients.

**Methods:** We conducted a multi-center retrospective study of positively confirmed COVID-19 patients from multiple hospitals in Louisiana. Demographics, medical history, comorbidities, clinical presentation, daily laboratory values, complications, and outcomes data were collected and analyzed. The primary outcome of interest was in-hospital mortality. Secondary outcomes were Intensive Care Unit (ICU) admission, risk of intubation, duration of mechanical ventilation, and length of hospital stay.

**Results:** A total of 502 COVID-19 patients (72 asthma and 430 non-asthma cohorts) were included in the study. The frequency of asthma in hospitalized cohorts was 14.3%, higher than the national prevalence of asthma (7.7%). Univariate analysis revealed that asthma patients were more likely to be obese (75% vs 54.2%, *p*=0.001), with higher frequency of intubation (40.3% vs 27.8%, *p* = 0.036), and required longer duration of hospitalization (15.1±12.5 vs 11.5±10.6, *p*=0.015). After adjustment, multivariable analysis showed that asthmatic patients were not associated with higher risk of ICU admission (OR=1.81, 95%CI=0.98-3.09, *p*=0.06), endotracheal intubation (OR=1.77, 95%CI=0.99-3.04, *p=*0.06) or complications (OR=1.37, 95%CI=0.82-2.31, *p*=0.23). Asthmatic patients were not associated with higher odds of prolonged hospital length of stay (OR=1.48, 95%CI=0.82-2.66, p=0.20) or with the duration of ICU stay (OR=0.76, 95%CI=0.28-2.02, *p*=0.58). Kaplan-Meier curve showed no significant difference in overall survival of the two groups (*p*=0.65).

**Conclusion:** Despite the increased prevalence of hospitalization in asthmatic COVID-19 patients compared to the general population, after adjustment for other variables, it was neither associated with increased severity nor worse outcomes.

## Introduction

Since its initial description in late 2019, the novel coronavirus (COVID-19) has become a global pandemic, overwhelming our healthcare systems and depleting resource stockpiles ^[1]^. In some pandemic hotspots, physicians have had to make difficult decisions regarding lifesaving interventions due to a lack of supplies such as ventilators ^[2]^. Further complicating management of this disease, COVID-19 has a broad clinical presentation. Patients can be asymptomatic carriers or have mild non-respiratory and respiratory symptoms. On the other hand, the virus can also present with severe pneumonia or lethal respiratory failure. Because of these severe outcomes, gaining an understanding of COVID-19’s disease course is essential to guiding treatment approaches and optimizing individual patient outcomes.

Preexisting comorbidities can have a dramatic impact on the course of a COVID-19 infection. It is therefore crucial to identify which comorbidities most contribute to severe infections, which may influence individual patient care plans ^[3]^. In efforts to better understand these relationships, the Centers for Disease Control and Prevention (CDC) is actively incorporating new and emerging data to update their treatment recommendations for patients with varying comorbid conditions. Currently, the CDC reports that asthma is present in about 17% of hospitalized COVID-19 patients, making it the fourth most prevalent comorbidity behind hypertension, obesity, and diabetes ^[4]^. Obese and diabetic patients have been categorized as high-risk, but there is still limited data regarding the impact of bronchial asthma on COVID-19 outcomes ^[5]^.

Because asthma is a chronic lung disease, it follows that asthmatics are at a greater risk for negative outcomes with a respiratory virus. However, there is a debate over the expected outcomes in COVID-19 infected patients with asthma. Some members of coronaviridiae family, that are associated with common cold, have been linked to asthma exacerbations ^[6]^. In contrast, Jackson et al. reported an observed protective relationship for asthma and atopic conditions in COVID-19 patients ^[7]^. In efforts to explain this phenomenon, it was proposed that patients with asthma and ectopic diseases may exhibit a lower concentration of respiratory tract angiotensin converting enzyme (ACE) receptors in patients. As has been widely reported, ACE receptors and specifically, ACE2, are known to be the host receptor site for SARS-CoV-2 ^[7,8]^.

Considering that more than 19 million adults in the Unites States have asthma ^[9]^, it is crucial to better characterize asthma in the setting of COVID-19. Therefore, we retrospectively evaluated hospitalized patients with laboratory confirmed SARS-CoV-2, focusing on the differing outcomes between asthmatic and non-asthmatic patients. The aim of this study was help further the understanding of asthma’s impact on COVID-19 outcomes, hopefully enabling physicians to tailor management and resource allocation in the response to the ongoing pandemic.

## Methodology

### Study population

Following Institutional Review Board (IRB) approval, data of hospitalized patients with laboratory-confirmed SARS-CoV-2 infection was gathered and entered into the web-based data collection platform, RedCap. Patients included in the study presented to either University Medical Center New Orleans (UMCNO), Tulane Medical Center (TMC), Saint Francis Medical Center Monroe (SFMC), or Our Lady of the Lake Regional Medical Center (OLOL) during the period between March 15 to June 9, 2020.

### Variables

A broad range of data was recorded including but not limited to the following: demographics, medical history, comorbidities, clinical presentation, daily laboratory values, complications, and outcomes. Patients were excluded if they were below 18 years of age or if they did not have recorded outcome data. Obesity was defined as body mass index (BMI) ≥30 Kg/m^2 [10]^. For severity assessment, CURB-65 score was estimated based on confusion status, respiratory rate≥30, blood pressure (systolic <90 mmHg or diastolic ≤60 mmHg), age ≥65 years, and blood urea nitrogen level >19 mg/dL (>7 mmol/L), and ranged from 0 to 5 ^[11]^. Quick Sequential Organ Failure Assessment (qSOFA) score was calculated using Glasgow coma score <15, respiratory rate ≥22, and systolic blood pressure ≤100, and was set to be a positive indicator for the poor outcome if ≥2 ^[12]^. Horowitz index (PaO_2_ / FIO_2_ ratio) was calculated as the ratio of arterial oxygen partial pressure (PaO_2_ in mmHg) to fractional inspired oxygen to determine a patient’s respiratory efficiency ^[13]^. The neutrophil-lymphocyte ratio (NLR), an indicator for poor outcome, was also estimated ^[14]^.

### Outcomes

A comparison between the asthmatic and non-asthmatic cohorts was performed. The primary outcome of interest was in-hospital mortality. Secondary outcomes were risk of Intensive Care Unit (ICU) admission, risk of endotracheal intubation, duration of mechanical ventilation, and length of hospital stay. Clinical diagnosis of complications was made using standard definitions such as the Risk, Injury, Failure, Loss of Kidney Function, and End-stage kidney disease (RIFLE) score for renal injury ^[15]^. Berlin criteria were also utilized to identify cases of acute respiratory distress syndrome (ARDS) ^[16]^

### Statistical analysis

Data management was performed in SAS 9.4 (*SAS Institute Inc*., *SAS 9*.*4, Cary, NC: SAS Institute Inc*., *2013*) and statistical analysis was carried out using SPSS 26.0 (*SPSS, Inc*., *Chicago, IL, USA*) and STATA 16.0 (*StataCorp. 2019. Stata Statistical Software: Release 16. College Station, TX: StataCorp LLC*). Continuous variables were described as medians and interquartile ranges, while categorical data were presented as frequencies and percentages. Chi-square or Fisher’s Exact tests were used for categorical variables, and Student’s t and Mann-Whitney U tests were applied for quantitative variables to examine the difference between asthmatic and non-asthmatics groups. Shapiro-Wilk test was used to test the normality of the continuous variables. Kaplan-Meier survival analysis and the Log Rank test was used to compare the in-hospital mortality in the two groups. Binary logistic regression analysis was used to assess the role of asthma comorbidity in the outcomes of COVID-19 disease. Age, gender, and obesity were adjusted in the model. Hosmer-Lemeshow test was used to assess goodness-of-fit. Results were reported as odds ratio (OR) and 95% confidence interval (CI) and two-sided p values below 0.05 were set as significant.

## Results

### Characteristics of the study population

A total of 502 confirmed positive COVID-19 patients were identified from multiple hospitals in Louisiana, including 72 bronchial asthma patients and 430 non-asthmatic controls. Their mean age was 60.6 ± 13.9 years and 60.8 ± 15.7 years respectively (*p* = 0.89). There was no observed gender difference; females accounted for 54.2% (39 patients) in the asthma group *versus* 52.0% (222 patients) in the non-asthma group (*p* = 0.79). The frequency of obesity among asthmatic patients (75%) was significantly higher than those without current asthma (54.2%), *p* = 0.001. On admission, asthma cohorts presented with greater respiratory rate (24.14 ± 7.03 vs 21.77 ± 7.20 breaths per minute, *p* = 0.015) and lower bicarbonate level (21.73 ± 5.41 vs 24.67 ± 3.57 mEq/L, *p* = 0.022). No other significant differences were found regarding clinical and laboratory features, **Table 1**.

**Table 1.** Baseline characteristics of COVID-19 patients at admission.

| Characteristics |  | Non-asthma (n=430) | Asthma (n=72) | P-value |
| --- | --- | --- | --- | --- |
| Age | Mean ± SD | 61.35 ± 15.64 | 57.69 ± 14.42 | 0.06 |
|  | 18-49 years | 91 (21.3) | 20 (27.8) | 0.20 |
|  | 50-64 years | 155 (36.2) | 29 (40.3) |  |
| | ≥ 65 years | 182 (42.5) | 23 (31.9) | |
| Sex | Female | 222 (52.0) | 39 (54.2) | 0.79 |
|  | Male | 205 (48.0) | 33 (45.8) |  |
| Race | African American | 325 (75.6) | 50 (69.4) | 0.09 |
|  | White | 66 (15.3) | 18 (25.0) |  |
|  | Not Reported | 39 (9.1) | 4 (5.6) |  |
| BMI, kg/m<sup>2</sup> | Mean ± SD | 32.90 ± 8.32 | 35.79 ± 9.43 | **0.034** |
| Smoking | None | 299 (69.5) | 47 (65.3) | 0.71 |
|  | Past smoker | 94 (21.9) | 17 (23.6) |  |
|  | Current smoker | 37 (8.6) | 8 (11.1) |  |
| Chief complaint | Shortness of breath | 231 (53.7) | 44 (61.1) | 0.25 |
|  | Fever | 94 (21.9) | 14 (19.4) | 0.76 |
|  | Cough | 93 (21.6) | 21 (29.2) | 0.17 |
|  | Flu-like symptoms | 26 (6) | 6 (8.3) | 0.44 |
|  | Fatigue | 28 (6.5) | 4 (5.6) | 0.71 |
|  | Chest pain | 10 (2.3) | 3 (4.2) | 0.41 |
|  | Altered mental status | 64 (14.9) | 6 (8.3) | 0.20 |
|  | Headache | 3 (0.7) | 2 (2.8) | 0.15 |
|  | Nausea, vomiting, diarrhea | 27 (6.3) | 4 (5.6) | 0.81 |
| Comorbidities | Hypertension | 306 (71.2) | 51 (70.8) | 0.95 |
|  | Diabetes | 183 (42.6) | 28 (38.9) | 0.60 |
|  | Chronic heart failure | 46 (10.7) | 4 (5.6) | 0.20 |
|  | Arrhythmia | 43 (10) | 5 (6.9) | 0.52 |
|  | COPD | 31 (7.2) | 5 (6.9) | 0.93 |
|  | Chronic kidney disease | 68 (15.8) | 8 (11.1) | 0.37 |
|  | Cancer | 47 (10.9) | 8 (11.1) | 0.96 |
|  | Coronary artery disease | 46 (10.7) | 3 (4.2) | 0.08 |
|  | Cerebrovascular disease | 34 (7.9) | 4 (5.6) | 0.63 |
|  | Obesity | 233 (54.2) | 54 (75) | **0.001** |
| Severity | qSOFA score | 0.69 ± 0.70 | 0.89 ± 0.72 | 0.06 |
| | CURB65 score | 1.47 ± 1.13 | 1.33 ± 1.20 | 0.41 |
| Orientation | Glasgow coma score | 13.59 ± 3.11 | 13.57 ± 3.57 | 0.98 |
| Vital signs | Temperature | 99.47 ± 1.67 | 99.75 ± 2.09 | 0.24 |
| | Pulse rate | 89.31 ± 19.41 | 93.78 ± 20.38 | 0.09 |
| | Systolic blood pressure | 125.52 ± 21.06 | 127.41 ± 20.20 | 0.51 |
| | Diastolic blood pressure | 73.66 ± 15.30 | 73.73 ± 13.44 | 0.97 |
| | Mean arterial pressure | 100.89 ± 18.52 | 101.75 ± 18.93 | 0.73 |
| | Respiratory rate | 21.77 ± 7.20 | 24.14 ± 7.03 | **0.015** |
| ABG findings | SaO<sub>2</sub> | 92.99 ± 7.80 | 93.59 ± 7.94 | 0.57 |
| | pH respiratory | 7.26 ± 1.01 | 7.39 ± 0.08 | 0.51 |
| | PaCO<sub>2</sub> | 39.24 ± 13.75 | 38.08 ± 10.53 | 0.68 |
| | PaO<sub>2</sub> | 89.67 ± 67.34 | 82.39 ± 78.09 | 0.62 |
| | HCO<sub>3</sub> | 24.67 ± 3.57 | 21.73 ± 5.41 | **0.022** |
| | FiO<sub>2</sub> (%) | 40.52 ± 29.58 | 43.18 ± 30.99 | 0.78 |
| | PaO<sub>2</sub>/FiO<sub>2</sub> ratio | 244.62 ± 111.31 | 236.00 ± 115.17 | 0.82 |
| Laboratory findings | White blood cells | 8.18 ± 5.70 | 7.95 ± 3.46 | 0.77 |
| | Hemoglobin | 12.11 ± 2.03 | 11.97 ± 2.31 | 0.64 |
|  | Hematocrit | 36.16 ± 5.91 | 36.70 ± 5.93 | 0.62 |
|  | Platelet count | 236.56 ± 104.9 | 246.15 ± 103. | 0.51 |
|  | Neutrophil count | 7.08 ± 9.91 | 6.20 ± 3.25 | 0.50 |
|  | Lymphocyte count | 1.33 ± 1.95 | 1.09 ± 0.43 | 0.34 |
|  | Neutrophil lymphocyte ratio | 7.97 ± 10.42 | 6.55 ± 4.51 | 0.30 |
|  | Serum sodium | 204.48 ± 902.78 | 137.24 ± 24.9 | 0.65 |
|  | Serum potassium | 4.09 ± 1.11 | 4.08 ± 0.86 | 0.98 |
|  | Serum chloride | 101.47 ± 8.85 | 101.82 ± 4.03 | 0.82 |
|  | Calcium corrected | 9.01 ± 0.66 | 8.99 ± 0.86 | 0.88 |
|  | Random blood sugar | 145.04 ± 86.29 | 155.97 ± 88.81 | 0.36 |
|  | Blood urea nitrogen | 26.50 ± 20.20 | 22.95 ± 20.89 | 0.20 |
|  | Serum creatinine | 1.82 ± 2.05 | 1.57 ± 1.88 | 0.37 |
|  | Albumin | 3.24 ± 0.56 | 3.36 ± 0.59 | 0.18 |
|  | Bilirubin | 0.64 ± 0.49 | 0.52 ± 0.19 | 0.23 |
|  | Alkaline phosphatase | 77.07 ± 45.08 | 65.54 ± 19.23 | 0.22 |
|  | AST | 48.59 ± 35.23 | 53.63 ± 38.84 | 0.53 |
|  | ALT | 33.01 ± 27.20 | 45.67 ± 35.96 | 0.05 |
|  | Anion gap | 12.22 ± 10.08 | 10.61 ± 2.68 | 0.50 |
|  | Lactic acid | 52.89 ± 104.58 | 1.75 ± 0.78 | 0.51 |
|  | Troponin | 3.04 ± 14.72 | 0.60 ± 0.94 | 0.78 |
|  | HbA1c | 7.51 ± 2.81 | 9.80 ± 4.81 | 0.33 |
|  | C-reactive protein | 48.21 ± 63.08 | 38.47 ± 58.98 | 0.64 |
|  | Procalcitonin | 11.49 ± 66.12 | 0.24 ± 0.22 | 0.51 |
|  | Ferritin | 987.87 ± 1,881.3 | 1,632.3 ± 3,293.5 | 0.26 |

Data are presented as mean and standard deviation or frequency and percentage. BMI: body mass index. SaO<sub>2</sub>: oxygen saturation, PaO<sub>2</sub>: partial pressure of oxygen, PaCO<sub>2</sub>: partial pressure of carbon dioxide, HCO<sub>3</sub>: bicarbonate, FiO<sub>2</sub>: Fraction of inspired oxygen, AST: Aspartate transaminase, ALT: alanine transaminase, HbA1c: glycosylated hemoglobin. Chi-square, Fisher's Exact, Student's t, or Mann-Whitney U tests were used. P-value at <0.05 was considered significant.

### Outcomes of asthma patients

Univariate analysis of COVID-19 outcomes revealed that asthma was significantly associated with higher rate of endotracheal intubation (40.3% vs 27.8%, *p* = 0.036), mechanical ventilation (both invasive and non-invasive) (70.7% vs 52.2%, *p* = 0.039), and longer hospital length of stay (15.14 ± 12.48 days vs 11.51 ± 10.58 days, *p* = 0.015). Asthma was not associated with a higher rate of Intensive Care Unit (ICU) admission (22.2% vs 14.9%, *p* = 0.12), acute respiratory distress syndrome (37.5% vs 30.9%, *p* = 0.27), or death (9.7% vs 13.5%, *p* = 0.45) among COVID-19 patients. No other clinical and laboratory findings were different between patients with and without asthma, **Table 2**.

**Table 2.** Comparison of the outcomes between asthmatic and non-asthmatic groups.

| Characteristics |  | Non-asthma<br>(n=423) | Asthma (n=72) | P value |
| --- | --- | --- | --- | --- |
| Procedures | Intubation | 118 (27.8) | 29 (40.3) | **0.036** |
|  | Extubation | 76 (64.4) | 25 (86.2) | **0.026** |
|  | Re-intubation | 10 (13.2) | 4 (16.0) | 0.74 |
|  | Mechanical ventilation | 109 (52.2) | 29 (70.7) | 0.039 |
|  | ICU admission | 63 (14.9) | 16 (22.2) | 0.12 |
| Days of events | Intubation days | 1.69 ± 2.53 | 2.62 ± 3.53 | 0.13 |
|  | Ventilation days | 2.05 ± 4.48 | 1.77 ± 2.70 | 0.72 |
|  | ICU LOS | 9.14 ± 7.22 | 9.00 ± 7.65 | 0.93 |
|  | ICU-free days | 9.77 ± 10.16 | 12.17 ± 10.68 | 0.10 |
|  | Total LOS | **11.51 ± 10.58** | **15.14 ± 12.48** | **0.015** |
|  | Time to death | 12.89 ± 8.79 | 14.71 ± 6.99 | 0.60 |
| Complications | None | 216 (50.2) | 30 (41.7) | 0.20 |
|  | One or more | 214 (49.8) | 42 (58.3) |  |
| Types of complications | ARDS | 133 (30.9) | 27 (37.5) | 0.27 |
|  | RF/AKI | 100 (23.3) | 16 (22.2) | 0.84 |
|  | Sepsis | 61 (14.2) | 14 (19.4) | 0.28 |
|  | Bacteremia | 28 (6.5) | 6 (8.3) | 0.61 |
| Current state | Still hospitalized | 7 (1.6) | 0 (0) | 0.60 |
|  | Closed cases | 423 (98.4) | 72 (100) |  |
| Mortality | Discharged alive | 366 (86.5) | 65 (90.3) | 0.45 |
|  | Died | 57 (13.5) | 7 (9.7) |  |
| Death location | ICU | 5 (8.8) | 1 (14.3) | 0.51 |
|  | Floor | 52 (91.2) | 6 (85.7) |  |

Seven non-asthmatic cases were still hospitalized thus was excluded from the analysis. ICU: intensive care unit, LOS: length of stay, ARDS: acute respiratory distress syndrome, RF/AKI: renal failure/acute kidney injury. Chi-square, Fisher's Exact, or Student's t were used. P-value at <0.05 was considered significant.

**Table 3.** Demographic and clinical outcomes of asthma and non-asthma patients stratified by obesity.

| Characteristics |  | Non-obese |  |  | Obese |  |  |
| --- | --- | --- | --- | --- | --- | --- | --- |
|  |  | Non-asthma (n=197) | Asthma (n=18) | P value | Non-asthma (n=233) | Asthma (n=54) | P-value |
| Age | Mean ± SD | 65.02 ± 16.32 | 59.50 ± 18.30 | 0.18 | 58.28 ± 14.38 | 57.09 ± 13.03 | 0.58 |
| Sex | Female | 80 (41) | 8 (44.4) | 0.81 | 135 (57.9) | 38 (71.7) | 0.09 |
|  | Male | 115 (59) | 10 (55.6) |  | 98 (42.1) | 15 (28.3) |  |
| Race | African American | 136 (69) | 15 (83.3) | 0.07 | 177 (76) | 47 (87) | 0.12 |
|  | White | 43 (21.8) | 0 (0) |  | 38 (16.3) | 3 (5.6) |  |
|  | Not Reported | 18 (9.1) | 3 (16.7) |  | 18 (7.7) | 4 (7.4) |  |
| Smoking | Active smokers | 20 (10.2) | 2 (11.1) | 0.98 | 17 (7.3) | 6 (11.1) | 0.48 |
| Severity | qSOFA | 0.65 ± 0.71 | 0.67 ± 0.65 | 0.95 | 0.71 ± 0.69 | 0.95 ± 0.74 | 0.05 |
| | CURB65 | 1.63 ± 1.16 | 1.58 ± 1.31 | 0.90 | 1.33 ± 1.08 | 1.25 ± 1.17 | 0.69 |
| | PF ratio | 269.47 ± 118.89 | 207.67 ± 67.00 | 0.39 | 229.33 ± 104.62 | 248.14 ± 133.52 | 0.67 |
| | Respiratory rate | 20.96 ± 6.98 | 20.94 ± 5.04 | 0.99 | 22.48 ± 7.33 | 25.21 ± 7.32 | **0.022** |
| Lab testing | Ferritin | 802.44 ± 771.40 | 3,192.06 ± 5,897.4 | 0.012 | 1,121.2 ± 2,379.5 | 923.37 ± 767.81 | 0.79 |
| | NLR | 8.37 ± 8.81 | 6.10 ± 4.33 | 0.34 | 7.61 ± 11.68 | 6.69 ± 4.60 | 0.61 |
| Procedures | Intubation | 37 (19.2) | 6 (33.3) | 0.22 | 81 (34.9) | 23 (42.6) | 0.29 |
|  | Extubation | 22 (59.5) | 5 (83.3) | 0.39 | 54 (66.7) | 20 (87) | 0.06 |
|  | Re-intubation | 3 (13.6) | 2 (40) | 0.22 | 7 (13) | 2 (10) | 0.73 |
|  | Mechanical ventilation | 35 (39.8) | 5 (62.5) | 0.27 | 74 (61.2) | 24 (72.7) | 0.22 |
|  | ICU admission | 42 (21.4) | 4 (22.2) | 0.93 | 77 (33.2) | 26 (48.1) | **0.042** |
| Days of events | Intubation days | 2.45 ± 3.30 | 2.00 ± 3.37 | 0.80 | 1.38 ± 2.07 | 2.73 ± 3.63 | **0.032** |
| | Ventilation days | 0.91 ± 2.85 | 0.63 ± 1.77 | 0.78 | 2.88 ± 5.23 | 2.11 ± 2.86 | 0.46 |
| | ICU LOS | 8.11 ± 7.17 | 7.50 ± 4.95 | 0.91 | 9.00 ± 6.84 | 7.50 ± 5.02 | 0.54 |
| | Total LOS | 11.72 ± 11.22 | 17.79 ± 10.82 | 0.05 | 11.34 ± 10.06 | 14.39 ± 12.91 | 0.08 |
| Complications | None | 107 (54.3) | 9 (50) | 0.81 | 109 (46.8) | 21 (38.9) | 0.36 |
|  | One or more | 90 (45.7) | 9 (50) |  | 124 (53.2) | 33 (61.1) |  |
| Types of complications | ARDS | 45 (22.8) | 5 (27.8) | 0.57 | 88 (37.8) | 22 (40.7) | 0.69 |
|  | RF/AKI | 44 (22.3) | 3 (16.7) | 0.77 | 56 (24) | 13 (24.1) | 1.00 |
|  | Sepsis | 28 (14.2) | 0 (0) | 0.14 | 33 (14.2) | 14 (25.9) | **0.042** |
|  | Bacteremia | 17 (8.6) | 0 (0) | 0.37 | 11 (4.7) | 6 (11.1) | 0.07 |
| Mortality | Discharged alive | 171 (89.1) | 17 (94.4) | 0.70 | 195 (84.4) | 48 (88.9) | 0.52 |
|  | Died | 21 (10.9) | 1 (5.6) |  | 36 (15.6) | 6 (11.1) |  |

PF ratio: PaO<sub>2</sub>: FiO<sub>2</sub> ratio, ICU: intensive care unit, LOS: length of stay, ARDS: acute respiratory distress syndrome, RF/AKI: renal failure/acute kidney injury.

Comparison stratified by obesity showed both asthmatic and non-asthmatic patients had overall similar demographic features, presenting manifestations, and comorbidities in each subgroup. However, clinical courses differed significantly between some groups. On comparison to non-asthmatic obese patients, obese asthmatic patients were more likely to develop sepsis (25.9% vs 14.2%, *p* = 0.042), had higher risk of ICU admission (48.1% vs 33.2%, *p* = 0.042), and required prolonged intubation (2.73 ± 3.63 days vs 1.38 ± 2.07, *p* = 0.032).**Impact of asthma comorbidity on COVID-19 outcomes**

Using multivariate regression analysis, bronchial asthma was not associated with higher risk of ICU admission (OR = 1.81, 95%CI = 0.98-3.09, *p* = 0.06) and endotracheal intubation (OR = 1.77, 95%CI = 0.99-3.04, *p =* 0.06). In addition, the presence of asthma was not associated with greater odds of complications (OR = 1.37, 95%CI = 0.82-2.31, *p* = 0.23), prolonged hospital length of stay (OR = 1.48, 95%CI = 0.82-2.66, p = 0.20) nor with the duration of ICU stay (OR = 0.76, 95%CI = 0.28-2.02, *p* = 0.58), **Figure 1A**. Kaplan-Meier curve showed no significant difference in overall survival of the two groups (*p* = 0.65), **Figure 1B**.

**Fig. 1.**
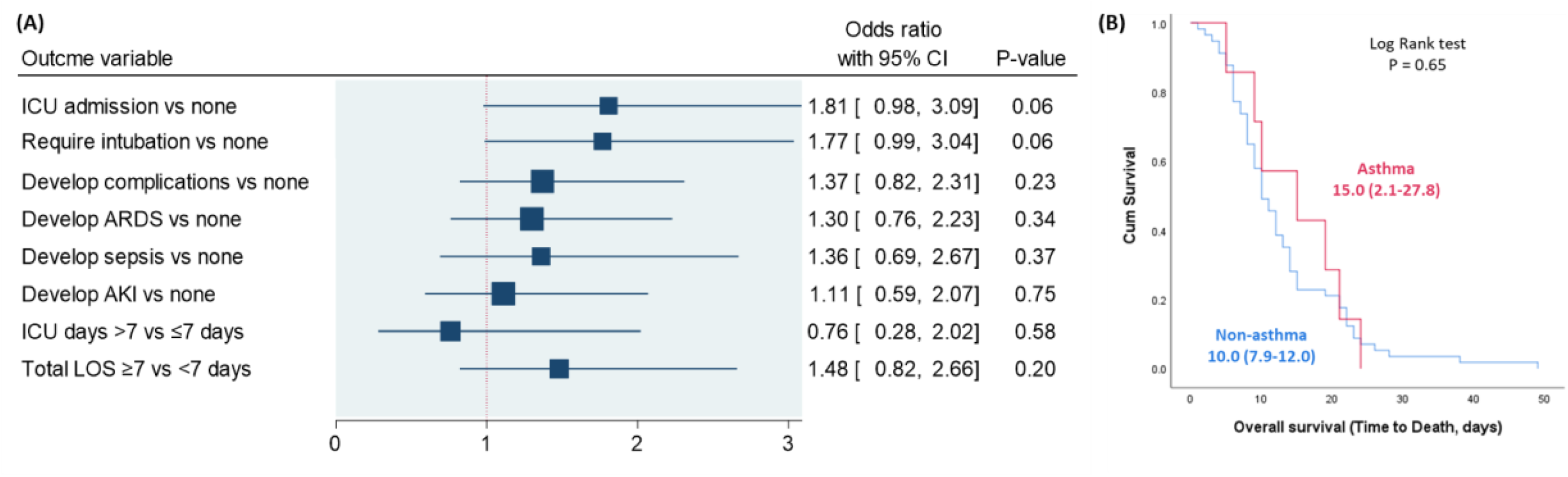
Impact of bronchial asthma on COVID-19 outcomes. (A) Binary regression analysis was performed. Odds ratio (OR) with 95% confidence interval (CI) was reported. The vertical reference line was set at 1. P-value at <0.05 was considered significant. Data were adjusted by age, gender, and obesity. ARDS: acute respiratory distress syndrome, AKI: acute kidney injury, ICU: intensive care unit, LOS: length of stay. (B) Kaplan-Meier survival curve comparing survival duration of asthma and non-asthma groups.

## Discussion

As the COVID-19 pandemic evolves, countries are considering policies to protect those at increased risk of severe disease. To the best of our knowledge, this is the first study evaluating the relationship between asthma and COVID-19 outcomes in hospitalized patients in Louisiana. In the 502 patients included, the prevalence of asthma was 14.3%. This rate is higher than the national prevalence of asthma reported by the CDC (7.7%), but in line with CDC reports on the prevalence of asthma in COVID-19 patients ^[17]^. This increased prevalence of asthma in COVID-19 patients may be attributable to increased disease awareness and concern to increased severity of the disease course among asthmatics.

Our initial univariate analysis of asthmatic patients showed significantly higher rates of endotracheal intubation and mechanical ventilation, as well as increased duration of hospital stay. Patients with asthma were also more likely to be obese, a finding consistent with prior studies ^[18,19]^. As has been well-documented and widely reported, obesity alone has been associated with poor clinical outcomes in hospitalized COVID-19 patients ^[4]^. Fittingly, in initial univariate analysis of obese asthmatic, there were significantly elevated rates of sepsis, ICU admission, and prolonged intubation. However, after multivariate adjustment, asthma comorbidity did not drive poor outcomes when it came to our primary and secondary endpoints of interest - ICU admission, intubation, mechanical ventilation, ARDS, and case fatality rate. Although these results may not be expected, there is a plausible mechanism, the downregulation of ACE2 receptors, that could hypothetically account for these observations ^[20]^.

Early data published by Jackson et al. suggested that patients with allergic asthma have decreased ACE2 expression in nasal and bronchial epithelial cells ^[7]^. The possible explanation for this observation is two-fold. First, it has been postulated that that type II immune modulation decreases the expression of the receptor for cellular entry of COVID-19 – ACE2 ^[22]^. Second, individuals with chronic respiratory conditions often use inhaled corticosteroids (ICS). The Severe Asthma Research Program (SARP) demonstrated that ICS use in asthmatic patients leads to decreased expression of ACE2 and TMPRSS2 - a host serine protease which primes host cells for entry via the spike protein ^[23]^. Additional *in vitro* studies have demonstrated that treatment with ciclesonide, an ICS used for suppression of asthma attacks, demonstrated viral suppression of SARS-CoV-2 ^[24]^. It must be remembered that these experimental studies occurred in non-COVID-19 patients, nonetheless, the results suggest further investigation is necessary to understand the possible protective role of type 2 inflammation in asthma and COVID-19. Based on these studies, it may be possible that ICS treatment is protective via viral suppression or ACE2/TMPRSS2 reduction. These protective effects may explain the lack of significant difference in outcomes among asthmatic and non-asthmatic patients.

The potential limitations of this study include self-reporting of asthma history and the inability to identify the severity of asthma. To prevent false reporting, each patient’s home medications were reviewed to confirm the diagnosis. Asthma severity may be assessed retrospectively based on symptoms or via risk assessment requiring measurement of airflow limitation using spirometry or Peak flow meter. However, these Aerosol Generating Procedures (AGPs) may put healthcare workers at an increased risk of coronavirus exposure.

The results of the current study show that while pre-existing asthma is more prevalent in our COVID-19 cohort than in the general population, after adjustment for other covariates, it was neither associated with disease severity nor negative outcomes. Therefore, although a seemingly poor prognosis, asthma does not imply a worse outcome as compared to non-asthmatics. This information can help physicians and researchers identify people with other underlying medical conditions at risk for more severe COVID-19 disease.

## Data Availability

Data is available upon request

## Acknowledgment

We thank numerous doctors, nurses, government, and civilians working together to fight against the SARS-CoV-2.

## Authors contribution

MHH, EAT, EK designed the study. ASA, MY, MO, NB, AZ, JR, AH, NA collected patients’ data. MHH performed data management. MHH, EAT performed statistical analyses. MH, EAT, EK contributed to data interpretation. MHH, EAT, NB, AZ, JR wrote the first draft. MHH, EAT drafted the final manuscript. All authors critically reviewed and approved the final version of the manuscript.

## References

1. Hamid S, Mir MY, Rohela GK. Novel coronavirus disease (COVID-19): a pandemic (epidemiology, pathogenesis and potential therapeutics). New Microbes New Infect. 2020;35:100679. Published 2020 Apr 14. doi:10.1016/j.nmni.2020.100679

2. Hick, J. L., D. Hanfling, M. K. Wynia, and A. T. Pavia. 2020. Duty to Plan: Health Care, Crisis Standards of Care, and Novel Coronavirus SARS-CoV-2. NAM Perspectives. Discussion paper. National Academy of Medicine. Washington, DC. https://doi.org/10.31478/202003b

3. Guan WJ, Liang WH, Zhao Y, et al. Comorbidity and its impact on 1590 patients with COVID-19 in China: a nationwide analysis. Eur Respir J. 2020;55(5):2000547. Published 2020 May 14. doi:10.1183/13993003.00547-2020

4. Garg S, Kim L, Whitaker M, et al. Hospitalization Rates and Characteristics of Patients Hospitalized with Laboratory-Confirmed Coronavirus Disease 2019 — COVID-NET, 14 States, March 1–30, 2020. MMWR Morb Mortal Wkly Rep 2020;69:458–464. DOI: http://dx.doi.org/10.15585/mmwr.mm6915e3

5. Centers for Disease Control and prevention. Coronavirus disease 2019 (COVID-19): are you at higher risk for severe illness? Atlanta, GA: US Department of Health and Human Services, CDC; 2020. https://www.cdc.gov/coronavirus/2019-ncov/specific-groups/high-risk-complications.html

6. Zheng XY, Xu YJ, Guan WJ, Lin LF. Regional, age and respiratory-secretion-specific prevalence of respiratory viruses associated with asthma exacerbation: a literature review. Arch Virol 2018; 163:845–53.

7. Jackson DJ, Busse WW, Bacharier LB, et al. Association of respiratory allergy, asthma, and expression of the SARS-CoV-2 receptor ACE2. J Allergy Clin Immunol. 2020;146(1):203-206.e3. doi:10.1016/j.jaci.2020.04.009

8. Hoffmann M, Kleine-Weber H, Schroeder S, et al. SARS-CoV-2 Cell Entry Depends on ACE2 and TMPRSS2 and Is Blocked by a Clinically Proven Protease Inhibitor. Cell. 2020;181(2):271-280.e8. doi:10.1016/j.cell.2020.02.052

9. Summary Health Statistics Tables for U.S. Adults: National Health Interview Survey, 2018, tables A-2b, A-2c. https://www.cdc.gov/nchs/fastats/asthma.htm (Last accessed July first)

10. Weir CB, Jan A. BMI classification percentile and cut off points. StatPearls. 2019.

11. George N, Elie-Turenne MC, Seethala RR, et al. External Validation of the qSOFA Score in Emergency Department Patients With Pneumonia. J Emerg Med. 2019;57(6):755–764. doi:10.1016/j.jemermed.2019.08.043

12. Su Y, Tu GW, Ju MJ, et al. Comparison of CRB-65 and quick sepsis-related organ failure assessment for predicting the need for intensive respiratory or vasopressor support in patients with COVID-19 [published online ahead of print, 2020 May 7]. J Infect. 2020;S0163-4453(20)30281-4. doi:10.1016/j.jinf.2020.05.007

13. Kao HC, Lai TY, Hung HL, et al. Sequential oxygenation index and organ dysfunction assessment within the first 3 days of mechanical ventilation predict the outcome of adult patients with severe acute respiratory failure. ScientificWorldJournal. 2013;2013:413216. doi:10.1155/2013/413216

14. Yang AP, Liu JP, Tao WQ, Li HM. The diagnostic and predictive role of NLR, d-NLR and PLR in COVID-19 patients. Int Immunopharmacol. 2020;84:106504. doi:10.1016/j.intimp.2020.106504

15. Kara I, Yildirim F, Kayacan E, Bilaloğlu B, Turkoglu M, Aygencel G. Importance of RIFLE (Risk, Injury, Failure, Loss, and End-Stage Renal Failure) and AKIN (Acute Kidney Injury Network) in Hemodialysis Initiation and Intensive Care Unit Mortality. Iran J Med Sci. 2017;42(4):397–403.

16. ARDS Definition Task Force, Ranieri VM, Rubenfeld GD, Thompson BT, Ferguson ND, Caldwell E, Fan E, Camporota L, Slutsky AS. Acute respiratory distress syndrome: the Berlin Definition. JAMA. 2012 Jun 20;307(23):2526–33.

17. Centers for Disease Control and prevention. Most Recent National Asthma Data. Source: 2018 National Health Interview Survey (NHIS) Data, Table 3-1 and Table 4-1. https://www.cdc.gov/asthma/most_recent_national_asthma_data.htm

18. Khalid F, Holguin F. A review of obesity and asthma across the life spac. J Asthma 2018; 55: 1286–300.

19. Peters U, Dixon AE, Forno E. Obesity and asthma. J Allergy Clin Immunol 2018; 141: 1169–79.

20. Finney, L. J. et al. (2020). Inhaled Corticosteroids Downregulate The SARS-Cov-2 Receptor ACE2 In COPD Through Suppression of Type I Interferon. bioRxiv preprint. doi: https://doi.org/10.1101/2020.06.13.149039. http://biorxiv.org/cgi/content/short/2020.06.13.149039

21. Chhiba KD, Patel GB, Vu THT, et al. Prevalence and characterization of asthma in hospitalized and non-hospitalized patients with COVID-19 [published online ahead of print, 2020 Jun 15]. J Allergy Clin Immunol. 2020;S0091-6749(20)30840-X. doi:10.1016/j.jaci.2020.06.010

22. Li W, Moore MJ, Vasilieva N, Sui J, Wong SK, Berne MA, et al., Angiotensin-converting enzyme 2 is a functional receptor for the SARS coronavirus. Nature 2003; 426(6965):450–4.

23. Peters MC, Sajuthi S, Deford P, et al. COVID-19-related Genes in Sputum Cells in Asthma. Relationship to Demographic Features and Corticosteroids. Am J Respir Crit Care Med. 2020;202(1):83–90. doi:10.1164/rccm.202003-0821OC

24. Matsuyama S, Kawase M, Nao N, Shirato K, Ujike M, Kamitani W, et al. The inhaled corticosterois ciclesonide blocks coronavirus RNA replication by targeting viral NSP15. BioRiv. 2020: DOI:10.1101/2020.03.11.987016.

